# Adoption of Guided Structured Reporting in Routine Radiological Practice: A Six-Week Multi-Site Implementation Study in the UAE

**DOI:** 10.64898/2026.05.20.26353646

**Authors:** Daniel Lorenz, Sven Jansen, Julius Knoche, Rüdiger Wolf-Sebottendorff, Hamzeh J. Awad, Igor Toker

**Affiliations:** Neo Q Quality in Imaging GmbH, Berlin, Germany; Honeycut & Peers GmbH, Bremen, Germany; Department of Physiotherapy, College of Pharmacy and Health Sciences, Ajman University, Ajman 346, United Arab Emirates; Center of Medical and Bio-Allied Health Sciences Research, Ajman University, Ajman 346, United Arab Emirates

**Keywords:** structured reporting, radiology, implementation, adoption, clinical workflow, artificial intelligence

## Abstract

**Background:** Guided structured reporting has been proposed to address the limited availability of structured data in radiology, yet empirical evidence on its real-world adoption across users and imaging modalities remains scarce.

**Objective:** To describe the adoption dynamics of a guided structured reporting system across multiple users and imaging modalities during a six-week implementation period.

**Methods:** Retrospective observational study at two public tertiary hospitals in Abu Dhabi, United Arab Emirates. A guided structured reporting system was deployed for computed tomography (CT), magnetic resonance imaging (MRI), and mammography. Seven radiologists participated. The primary outcome was active in-software reporting time, recorded via system logs of mouse and keyboard interaction. Temporal trends in median reporting time per modality and individual user trajectories were analysed descriptively. After predefined data cleaning, 126 reports were included (84 CT, 27 MRI, 15 mammography).

**Results:** Active in-software reporting time decreased across all modalities. Median reporting time fell from 130 s to 56 s for CT, from 383 s to 60 s for MRI, and from 126 s to 46 s for mammography (week 1 to week 6). Individual trajectories showed similar patterns, with the largest reductions during the early implementation phase. Subgroup analyses were limited by small sample sizes.

**Conclusions:** Guided structured reporting was integrated into routine clinical workflows with temporal reductions in active reporting time across users and modalities, providing empirical evidence on the feasibility of workflow-integrated structured reporting in radiological practice.

**Plain Language Summary:** *What is the problem?:* Radiology reports are often written as free text, which makes them difficult to use for research, quality improvement, or training artificial intelligence systems. Structured reporting tools aim to standardise how radiologists document their findings, but we know little about how well these tools work in everyday hospital practice — especially across different types of medical imaging and different users.

*What did we do?:* We introduced a guided structured reporting system at two hospitals in Abu Dhabi, United Arab Emirates. Over six weeks, seven radiologists used the software during their normal working shifts to create reports for CT scans, MRI examinations, and mammograms. The software guides the user through a structured template and records how long they spend actively interacting with it. We tracked these interaction times to see how quickly radiologists became familiar with the system.

*What did we find?:* Across all three types of imaging, the time radiologists spent interacting with the software decreased over the six-week period. For CT, the median active time dropped from about two minutes to under one minute. For MRI, times decreased from over six minutes to one minute. Individual radiologists showed similar patterns: most of the improvement happened in the first few weeks, after which reporting times stabilised. The proportion of reports exported to the clinical system also increased over time, suggesting growing integration into routine workflows.

*What does this mean?:* Our findings suggest that guided structured reporting can be adopted in everyday hospital practice without major disruption. Radiologists became more efficient with the system over a relatively short period. However, this study measured only the time spent within the software, not overall report quality or clinical outcomes. Further research is needed to determine whether structured reporting also improves data quality or supports downstream applications such as clinical decision-making tools.

## Introduction

Artificial intelligence (AI) has achieved high levels of diagnostic performance in controlled evaluation settings in radiology [1, 2], yet its impact on routine clinical practice remains limited [3, 4]. This reflects a well-recognised challenge in digital health: technologies that demonstrate strong performance in pilot environments often fail to achieve sustained adoption in routine clinical workflows [5, 6]. Factors such as workflow integration, usability, and system compatibility frequently determine whether innovations translate into clinical impact, independent of their technical capabilities.

A key barrier is the state of clinical data itself. Many AI applications rely on structured, consistent, or well-annotated data, but everyday radiological reporting is still largely based on unstructured or variably structured formats [7–10]. As a result, the data required to support scalable AI applications are often not readily available during routine care [11, 12]. Current research has predominantly focused on algorithm development and validation, while less attention has been directed toward the upstream conditions required for effective deployment—particularly whether structured data generation can be implemented and sustained within routine clinical workflows [13, 14].

Structured and guided reporting systems have been introduced as a potential approach to address this challenge by standardising report content and format during the reporting process. However, empirical evidence on their real-world adoption over time remains limited, especially across multiple users and imaging modalities [9, 15].

A recent Nature editorial on proportional evidence in digital health emphasises that claims of clinical impact should be matched by appropriate evidence: implementation studies for workflow integration claims, and prospective comparative evaluations for outcome or efficiency claims [16]. This study responds to the former by examining the adoption of a guided structured reporting system in routine clinical practice over a six-week period. Rather than evaluating diagnostic performance or downstream outcomes, the focus lies on adoption dynamics and workflow integration, assessed through temporal changes in active in-software reporting time across multiple users and modalities.

## Methods

### Study Design

This study was designed as a retrospective observational case study and is reported in accordance with the STROBE guidelines for observational studies (Supplementary Checklist) of a six-week implementation pilot of guided structured reporting in a real-world clinical setting. The objective was to describe the implementation process and adoption of a structured reporting system during routine radiological practice within an operational hospital environment. No control group was included, and the analysis focuses exclusively on observed patterns within the implementation period.

### Setting and Participants

The study was conducted at two public tertiary hospitals (732 and 586 beds, respectively) in Abu Dhabi, United Arab Emirates. The implementation took place in routine clinical practice without experimental conditions.

A total of seven radiologists participated in the pilot. These included board-certified radiologists involved in daily reporting activities. One radiologist used the reporting system across two separate technical instances - one on-site within the hospital network and one remotely via a web-based environment. As both accounts corresponded to the same individual, these were consolidated and treated as a single participant for the purpose of analysis.

All participants were pseudonymised in the source data using system-generated unique identifiers and are referred to by generic labels (Radiologist A–G) throughout this manuscript.

### Intervention (Guided Structured Reporting)

The intervention consisted of the implementation of a guided structured reporting system (RadioReport®, Neo Q Quality in Imaging GmbH) into routine clinical workflows. The system enables radiologists to create structured radiology reports through a guided interface, using predefined templates and guided, decision-tree-based navigation across anatomical regions and findings.

Radiologists used the system as part of their regular reporting activities during the pilot period. The software was integrated into the existing reporting environment and applied in routine clinical practice as part of a structured implementation pilot, without restrictions on case selection beyond standard operational workflows.

The intervention focused on the use of guided reporting for routine examinations across multiple imaging modalities, including computed tomography (CT), magnetic resonance imaging (MRI), and mammography.

### Implementation and Training

The implementation followed a structured hybrid training and rollout model combining online and on-site components. Prior to the start of the pilot, all participating radiologists completed an initial online training phase consisting of a two-hour introductory session and a two-hour web-based course. This phase included an introduction to the software, demonstration of the reporting workflow, and access to an e-learning platform with case-based exercises.

The on-site implementation was conducted during the first week of the pilot. This included a four-hour group training session followed by individualized 1:1 training sessions with each radiologist. During these sessions, participants were guided through the reporting workflow and performed supervised reporting, including the completion of five reports per radiologist under instructor supervision.

Following the initial training phase, radiologists began using the system in routine clinical practice. During the second week, on-site support was provided by the implementation team to assist with early adoption. In weeks three to five, continued system use was supported through remote assistance and ongoing user feedback. The final week of the pilot was dedicated to data collection and evaluation.

Training was delivered using a combination of standardized components (online modules and group sessions) and individualized 1:1 sessions tailored to the experience and needs of each participant.

### Outcome Definition

The primary outcome of the study was the active in-software reporting time. This metric represents the duration of user interaction within the reporting system, as recorded by system logs of mouse and keyboard interaction. It reflects the portion of the reporting process directly attributable to software use and does not include time spent on image interpretation or other activities performed outside the system.

Active in-software reporting time serves as a measure of workflow engagement with the structured reporting interface, not as a proxy for overall reporting efficiency or clinical turnaround time. A reduction in this metric may reflect increasing familiarity with the software interface and template navigation, but does not necessarily indicate faster overall report completion. Activities such as image review, clinical reasoning, dictation, and report verification occur outside the software and are not captured by this metric.

### KPI Framework

Pre-defined efficiency key performance indicators (KPIs) were established as part of the institutional pilot agreement, specifying target reporting times per modality (CT, MRI, mammography) and minimum on-target percentages. As the baseline measurement methods used by the institution could not be independently verified and may reflect a different metric (e.g., total dictation time rather than active in-software time), KPI achievement is reported for contractual context only (see supplement) and does not constitute the primary analysis of this study.

### Data Sources and Data Processing

Data were collected from the RadioReport® system deployed at two public tertiary hospitals during the six-week pilot. Each record in the dataset corresponds to a single radiological report generated within the system. Seven radiologists used the platform during routine clinical shifts; one radiologist additionally used the system remotely via a web-based instance for evening reporting sessions. All instances shared identical software, templates, and module configuration. A sensitivity analysis excluding all remote-instance data confirmed the robustness of the findings (Supplementary Table S4).

All data were processed using structured export files containing reporting activity metrics and timestamps. The consolidated dataset served as the basis for all subsequent analyses.

### Data Cleaning

Prior to analysis, the dataset underwent a predefined data cleaning process. Entries with missing or non-valid timestamps or reporting time values were excluded.

To reduce the impact of non-representative or technically implausible values, entries with an active reporting time below 20 seconds were removed, as this duration is insufficient to navigate the minimum required template fields and likely reflects incomplete or aborted reporting sessions. Upper outliers were excluded per modality using the standard interquartile range (IQR) method (values above Q3 + 1.5 × IQR). The impact of these thresholds on the results was assessed through sensitivity analyses using alternative cut-offs (no lower bound; 2 × IQR upper bound), reported in the supplement.

This approach was applied consistently across the dataset to ensure comparability and robustness of the descriptive analyses.

### Statistical Analysis

Descriptive statistics included median active reporting time with interquartile range (IQR) per modality and week. To quantify uncertainty, 95% bias-corrected and accelerated (BCa) bootstrap confidence intervals were computed using 10,000 resamples. Temporal trends were assessed using Spearman rank correlation between chronological report sequence number and active reporting time per modality. A linear mixed-effects model was fitted with log-transformed active reporting time as the dependent variable, POC week and modality as fixed effects, and a random intercept for radiologist, to account for repeated measures within users. Five pre-specified sensitivity analyses were conducted to assess the robustness of observed trends under alternative analytical choices (Supplementary Table S4). Bootstrap confidence intervals, Spearman correlations, and mixed-effects model results are reported in Supplementary Tables S1–S3. All analyses were performed using Python (scipy, statsmodels).

To characterize the learning curve, a rolling average over consecutive reports was calculated per user using a window of ten reports, chosen to balance noise reduction against temporal resolution given the available sample size. In addition, a linear trend line was fitted to the data to visualize overall changes in reporting time over the observation period.

### Ethical and Data Protection Considerations

This study analysed system-generated performance metadata (active in-software reporting time, session timestamps, and interaction counts) that did not include patient-identifiable information or clinical report content. Formal ethics approval was not required for this study, as it involved only anonymised operational performance metrics from a software implementation. No patient data, human tissue, or personally identifiable information was collected or processed. Informed consent was not applicable.

All radiologists were pseudonymised in the source data via system-generated unique identifiers; no personally identifiable information was included in the analytical dataset. Data processing was performed within the institutional IT environment in accordance with applicable data protection regulations and internal governance policies.

The data used in this study are not publicly available due to institutional and data protection restrictions but may be available from the corresponding author upon reasonable request and with permission of the respective institution.

## Results

### Dataset Overview

The final analysis dataset consisted of reporting activity collected over a six-week implementation period. Following data cleaning, a total of 126 reports were included in the analysis.

The dataset comprised reporting activity from seven radiologists working across two hospital sites within a public hospital network, including one radiologist contributing data via a separate system instance used remotely. These data were merged into a unified dataset reflecting both on-site and remote system usage.

Examinations were categorized by imaging modality based on module names, including computed tomography (CT), magnetic resonance imaging (MRI), and mammography. Of the included reports, 84 were classified as CT, 27 as MRI, and 15 as mammography. The full module-to-modality classification is reported in the Supplementary Modality Mapping Table.

All data included timestamped records of reporting activity and corresponding active in-software reporting times, forming the basis for subsequent temporal and descriptive analyses.

Beyond reporting time, additional indicators of system adoption were derived from the source data. All 126 reports were generated to completion (completion rate 100%). Of these, 91 (72.2%) were exported to the clipboard or clinical system. The export rate increased markedly over the course of the pilot, rising from 32% in week 1 to 94% in week 5, suggesting growing integration of structured reports into the clinical workflow. The majority of sessions (113/126, 89.7%) were closed on the final page of the guided reporting workflow, which progresses through sequential sections from clinical context to findings and conclusions, while 13 (10.3%) were closed on an earlier section, suggesting premature session termination. Median total session time was 427 s (IQR 184–1027), of which a median of 104 s (39.5%) was active in-software reporting time, indicating that the majority of session time was spent on activities outside direct software interaction (e.g., image review, clinical reasoning). Adoption metrics stratified by week and radiologist are reported in Supplementary Table S7.

### Temporal Development of Reporting Time by Modality

The temporal development of active in-software reporting time across the six-week implementation period is shown in Figure 1. Reporting performance is presented as the median active reporting time per week, stratified by imaging modality (CT, MRI, and mammography).

**Figure 1.**
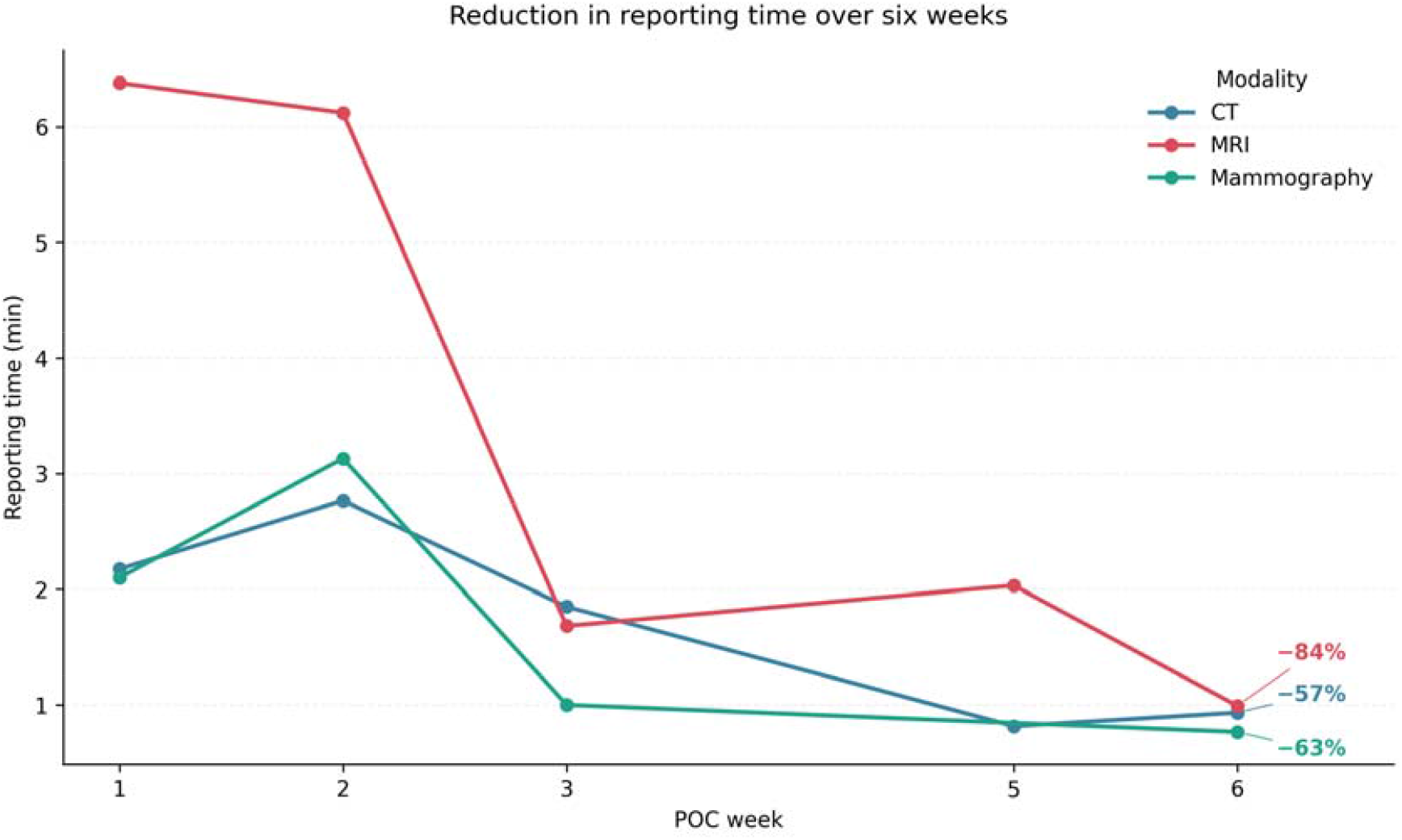
Median active in-software reporting time per week, stratified by imaging modality (CT, MRI, mammography). N = 126 reports from 7 radiologists.

Across all modalities, a reduction in median reporting time was observed over the course of the pilot. Weekly medians with interquartile ranges are summarised in Table 1. For CT (n = 84), median reporting time fell from 130 s (IQR 93–193) in week 1 to 56 s (IQR 33–102) in week 6, representing a 57% reduction. MRI (n = 27) showed the largest absolute decrease, from 383 s (IQR 321–442) in week 1 to 60 s (IQR 54–65) in week 6, a reduction of 84%. Mammography (n = 15) decreased from 126 s (IQR 94–238) in week 1 to 46 s in the final observation week, a reduction of 63%. Bootstrap confidence intervals for weekly medians are reported in Supplementary Table S1. Temporal trends for individual modalities were limited by small subgroup sizes, particularly for MRI (n = 2 in week 6) and mammography (n = 1 in week 6).

**Table 1.** Median active in-software reporting time (seconds) with interquartile range (IQR) per imaging modality and POC week. No reports were recorded in week 4. Mammography had no reports in week 5.

| Modality | Week | n | Median (s) | IQR |
| --- | --- | --- | --- | --- |
| CT | 1 | 12 | 130 | 93–193 |
| CT | 2 | 15 | 166 | 91–258 |
| CT | 3 | 21 | 111 | 50–171 |
| CT | 5 | 25 | 49 | 29–93 |
| CT | 6 | 11 | 56 | 33–102 |
| **CT** | **Overall** | **84** | **92** | **46–170** |
| MRI | 1 | 8 | 383 | 321–442 |
| MRI | 2 | 4 | 368 | 266–473 |
| MRI | 3 | 6 | 101 | 94–113 |
| MRI | 5 | 7 | 122 | 78–156 |
| MRI | 6 | 2 | 60 | 54–65 |
| **MRI** | **Overall** | **27** | **168** | **95–350** |
| Mammography | 1 | 5 | 126 | 94–238 |
| Mammography | 2 | 6 | 188 | 86–268 |
| Mammography | 3 | 3 | 60 | 55–134 |
| Mammography | 6 | 1 | 46 | n/a |
| **Mammography** | **Overall** | **15** | **126** | **58–224** |

While the magnitude and temporal pattern of change differed between modalities, all three showed lower median reporting times in later stages of the implementation period compared to the initial weeks. Overall, the results indicate a temporal reduction in median active reporting time across modalities over the six-week period.

Spearman rank correlation confirmed a statistically significant monotonic decrease in reporting time for all three modalities (CT: rho = -0.40, p < 0.001; MRI: rho = -0.78, p < 0.001; mammography: rho = -0.55, p = 0.034). A linear mixed-effects model with random intercept for radiologist estimated a 23.2% reduction in active reporting time per additional week (coefficient for week: -0.264, 95% CI -0.352 to -0.177, p < 0.001), after adjusting for modality and between-user variability. Sensitivity analyses using alternative outlier thresholds, excluding the highest-volume user, excluding remote-instance data, and excluding initial training reports confirmed the direction and general pattern of the trend, with expected loss of statistical significance in reduced subsamples (Supplementary Table S4).

### Individual Adoption Patterns

Individual reporting patterns over time are illustrated in Figure 2, which shows the temporal development of active in-software reporting time for the radiologist with the highest report volume (n = 71, 56% of total reports) across the observation period.

**Figure 2.**
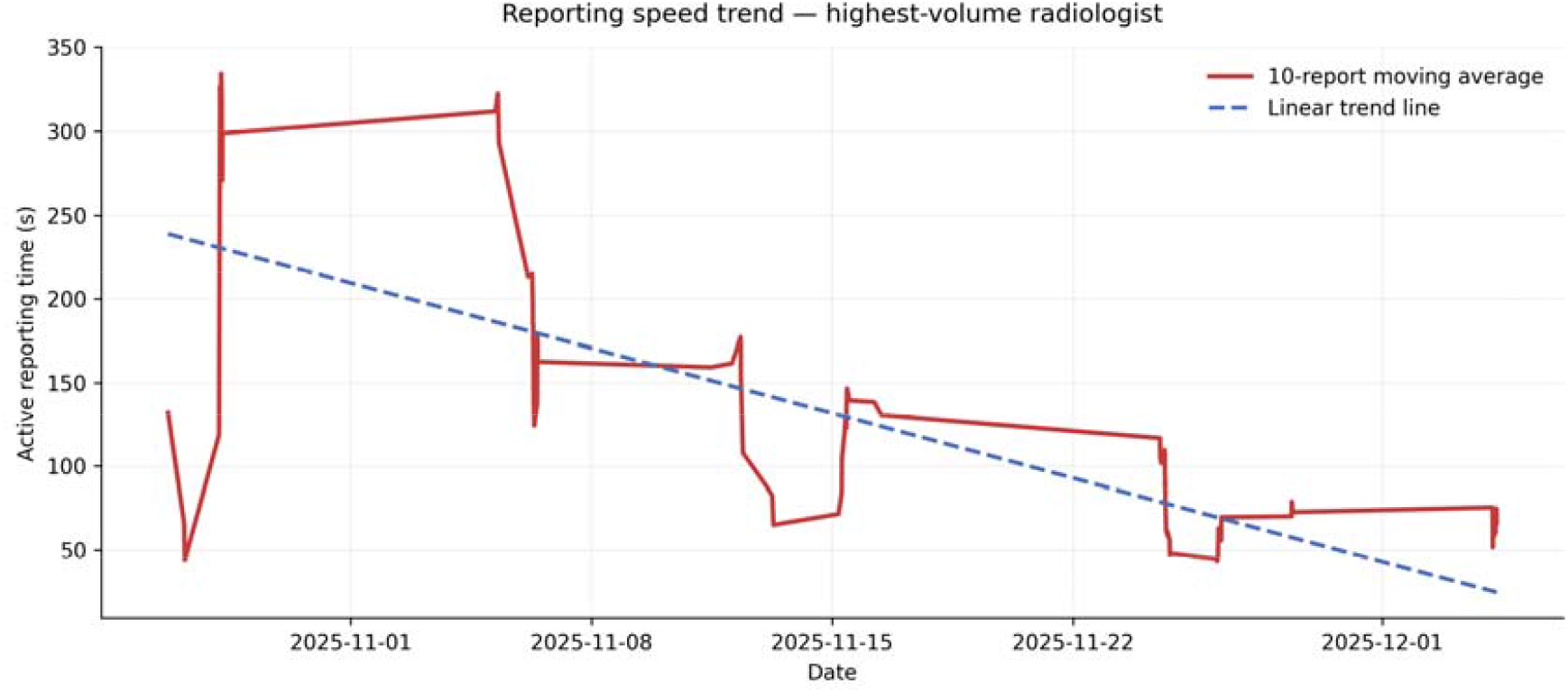
Individual trajectory of active in-software reporting time for the highest-volume radiologist (n = 71 reports). Dots represent individual reports; the solid line shows a 10-report rolling average; the dashed line indicates the fitted linear trend.1.

The individual trend shows a progressive reduction in reporting time over the course of the pilot. Early reports are characterized by higher and more variable reporting times, followed by a gradual stabilization and reduction in later stages. The rolling average indicates a consistent downward trend, while the fitted linear trend line suggests an overall decrease in reporting time across the observation period.

Despite variability at the level of individual reports, the overall trajectory remains stable, with no sustained increase in reporting time in later phases. The observed pattern is consistent with the temporal trends shown at the modality level, indicating that reductions in reporting time are not solely driven by aggregation effects but are also reflected at the level of individual users.

To assess whether the observed temporal trends generalised beyond the highest-volume user, individual learning curves for all seven radiologists are presented in Figure 3. Despite substantial variation in report volume (range: 4–71 reports per radiologist), a downward trend in median reporting time was visible for most participants with sufficient data points. Radiologists A, B, and F showed clear reductions across modalities, while radiologists with fewer reports (D, G) had too few observations per week for meaningful trend assessment. The consistency of temporal patterns across individual users supports the interpretation that the observed reductions reflect a general adoption effect rather than being driven by a single high-volume contributor.

**Figure 3.**
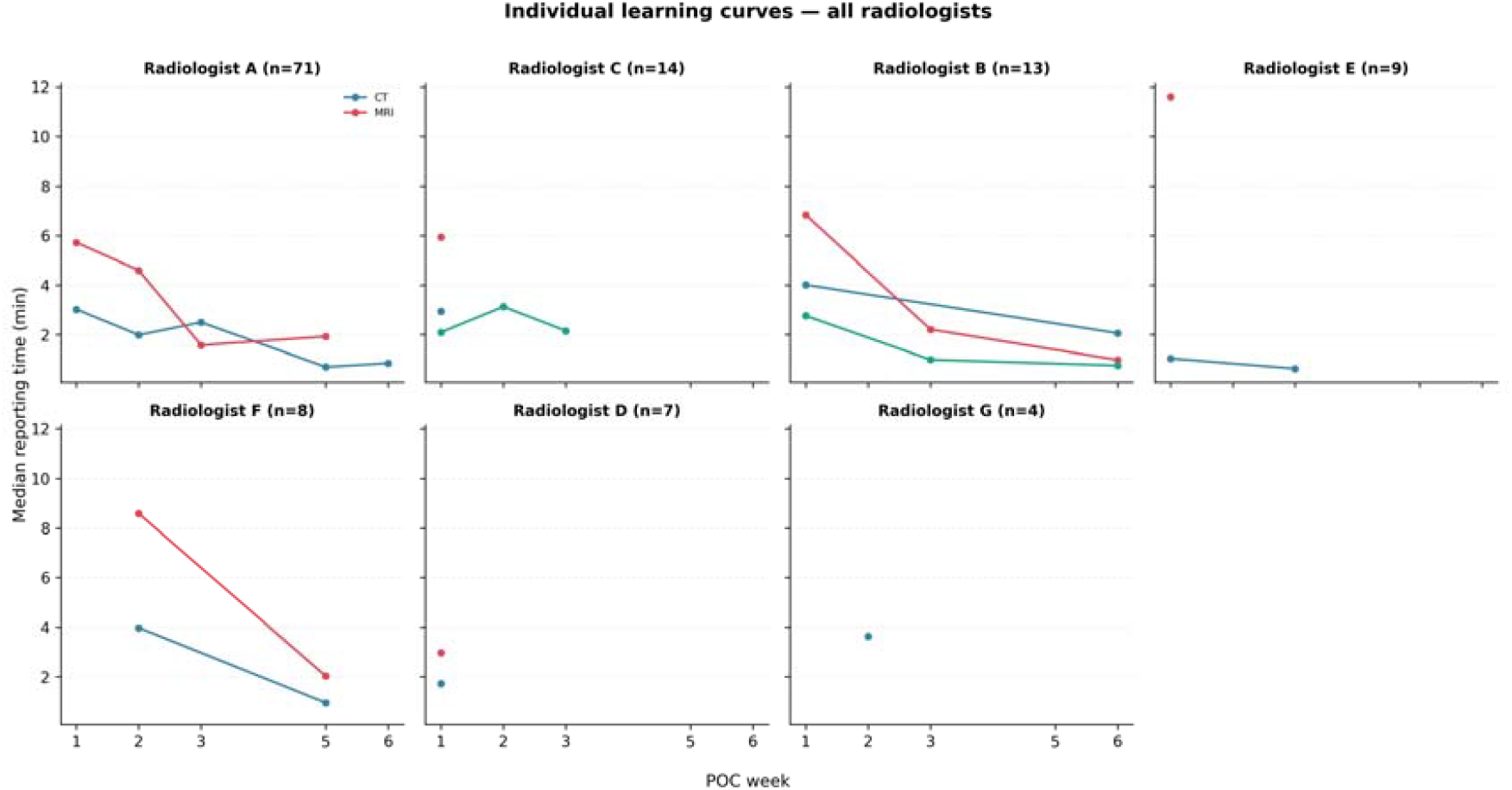
Individual learning curves for all seven radiologists, showing median active in-software reporting time per week, stratified by imaging modality. Panel titles indicate anonymised radiologist label and total report count. Note the shared y-axis across panels.

## Discussion

### Key Findings

This study observed a temporal reduction in active in-software reporting time over a six-week implementation period of guided structured reporting in a real-world clinical setting. Temporal trends across modalities showed a progressive decrease in median reporting time, with the most pronounced reductions occurring during the early phase of system use, followed by stabilization in later weeks. These patterns were observed both at the modality level and at the level of individual users, suggesting that the observed patterns are not solely driven by aggregation.

Beyond temporal trends in reporting time, secondary adoption indicators—including a completion rate of 100% and an increase in export rate from 32% to 94%—suggest progressive clinician acceptance of the structured reporting workflow over the observation period.

### Interpretation in Context of Implementation

The observed temporal patterns are consistent with a typical adoption process in digital health implementations [17, 18]. This pattern aligns with broader observations in digital health adoption, where the introduction of new technologies not only affects workflow efficiency but also exposes underlying structural dependencies within clinical systems. Initial variability and higher reporting times in early phases were followed by progressive stabilization and reduction, suggesting adaptation to the structured reporting workflow.

Notably, this implementation was conducted in a healthcare system outside Europe, where structured reporting has been less widely studied. The observed adoption dynamics—including the temporal pattern and magnitude of reporting time reductions—suggest that guided structured reporting may generalise across diverse operational and regulatory healthcare environments, though further comparative studies are needed to confirm this.

Importantly, the results reflect changes in active in-software reporting time rather than total report turnaround time. The metric captures direct user interaction within the reporting system and therefore provides a focused view on workflow engagement rather than end-to-end clinical efficiency.

### Implications within a proportional-evidence framework

A recent Nature editorial has called for proportional evidence in digital health, arguing that claims of clinical impact should be matched by the type of evidence appropriate to the claim: implementation studies for workflow integration, and prospective comparative evaluations for outcome or efficiency claims [16]. The present study aligns with the former tier: it provides pilot implementation evidence that guided structured reporting can be integrated into routine clinical workflows without introducing delay or disruption, but it does not claim efficiency gains or improved clinical outcomes. Beyond workflow integration, the adoption of structured reporting may carry implications for clinical data infrastructure. Compared to free-text reporting, structured templates reduce variability and generate standardised outputs that are prerequisites for many AI applications [19–23]. By embedding standardisation directly into the reporting process, structured reporting may reduce the need for retrospective data curation [24, 25]. Whether the data generated during this implementation are of sufficient quality and consistency for downstream applications warrants dedicated investigation.

### Strengths and Limitations

This study is based on real-world implementation data from multiple clinical sites and users over a defined pilot period. The analysis captures both temporal trends and individual usage patterns, providing a multi-level perspective on system adoption. The use of objective, system-derived reporting time metrics reduces reliance on subjective assessments and allows for reproducible analysis.

Several limitations should be considered. First, the study evaluates active in-software reporting time rather than total reporting turnaround time, and therefore does not capture all aspects of reporting efficiency. A reduction in active time may reflect increasing familiarity with template navigation rather than faster overall report completion. Second, the first week of the pilot included supervised training sessions (five reports per radiologist); we cannot fully disentangle the relative contributions of initial training, software familiarisation, and genuine adoption to the observed time reductions. A sensitivity analysis excluding the first five reports per user confirmed the direction of the trend for CT and MRI, though with reduced statistical significance (Supplementary Table S4). The distribution of examination types across weeks showed no systematic shift toward less complex examinations (Supplementary Table S5). Third, the observation period is limited to six weeks, which may not reflect long-term usage patterns, and no reports were recorded in week 4 across any modality. Fourth, time-of-day patterns differed across weeks: week 5 included a substantial proportion of evening reports from a single remote user, which were associated with shorter reporting times. This temporal clustering may partly explain the low median values observed in later weeks (Supplementary Table S6). Fifth, the analysis is based on a limited number of users (n = 7) within a pilot setting; one radiologist contributed 56% of all reports, and subgroup sizes for MRI and mammography were small (n = 27 and n = 15, respectively), limiting the precision of modality-specific estimates. Sixth, we did not have access to the total number of eligible examinations per department, precluding calculation of an adoption rate in the strict sense. Finally, no pre-implementation baseline using the same metric was available; all comparisons are within-pilot temporal trends.

## Conclusion

Guided structured reporting was adopted in routine clinical workflows over a six-week implementation period, with temporal reductions in active in-software reporting time observed across multiple imaging modalities and users. These findings provide empirical evidence on the feasibility of workflow-integrated structured reporting in routine radiological practice. The adoption of structured reporting may contribute to generating standardised datasets relevant for future clinical AI applications, though this was not directly evaluated in the present study.

## Declarations

### Contributors

DL conceived the study, performed the data analysis, and drafted the manuscript. SJ contributed to study conceptualisation and critical revision of the manuscript. JK provided domain expertise and contributed to critical revision of the content. RWS coordinated the on-site implementation and project management during the pilot. HJA supervised the ethics process and contributed to the critical revision of the manuscript. IT contributed to conceptualisation, supervised the study, and critically revised the manuscript for intellectual content. IT is the guarantor of the article and accepts full responsibility for the integrity of the work as a whole. All authors had access to the raw data, reviewed the final manuscript, and approved the version submitted for publication.

### Use of AI-assisted tools

We used Anthropic’s Claude (Claude Code, Opus model) and OpenAI’s ChatGPT to support the drafting and refinement of the manuscript. The AI tools were employed as writing assistants to improve clarity, structure, and language. Statistical analyses were implemented, verified, and interpreted by the authors. No AI was used for data generation or interpretation of scientific results. The final text was reviewed and edited by the authors to ensure accuracy, originality, and appropriateness for publication.

### Declaration of conflicting interest

SJ is the CEO of Neo Q Quality in Imaging GmbH. Three further authors (DL, RWS, IT) are employees of the same company, the company that develops RadioReport®, the guided structured reporting software evaluated in this study. JK is an employee of Honeycut & Peers GmbH, a healthcare consulting firm that has worked with Neo Q. HJA serves as a Digital Health Advisor to Neo Q.

Neo Q provided the RadioReport® software and technical support for the implementation pilot. The data analysis was performed by DL. All authors had full access to the study data and the corresponding author (DL) had final responsibility for the decision to submit for publication. Neo Q had no veto right over the publication, and the study was conducted and reported independently by the authors.

### Funding

The authors received no specific grant for this research from any funding agency in the public, commercial, or not-for-profit sectors. The implementation pilot was conducted as part of a contractual agreement between Neo Q Quality in Imaging GmbH and Pure Health (Abu Dhabi). Neo Q provided the software and on-site implementation support; no additional financial resources were allocated for the research or publication.

### Ethical considerations

This study analysed system-generated performance metadata (active in-software reporting time, session timestamps, and interaction counts) that did not include patient-identifiable information or clinical report content. All participating radiologists were pseudonymised using system-generated unique identifiers; no personally identifiable information was included in the analytical dataset.

Formal ethics approval was not required for this study, as it involved only anonymised operational performance metrics from a software implementation. No patient data, human tissue, or personally identifiable information was collected or processed.

### Data availability

The anonymised performance data supporting the findings of this study are available from the corresponding author upon reasonable request and with permission of the participating institution. Raw data cannot be publicly shared due to contractual confidentiality agreements with Pure Health.

### CRediT Author Contributions

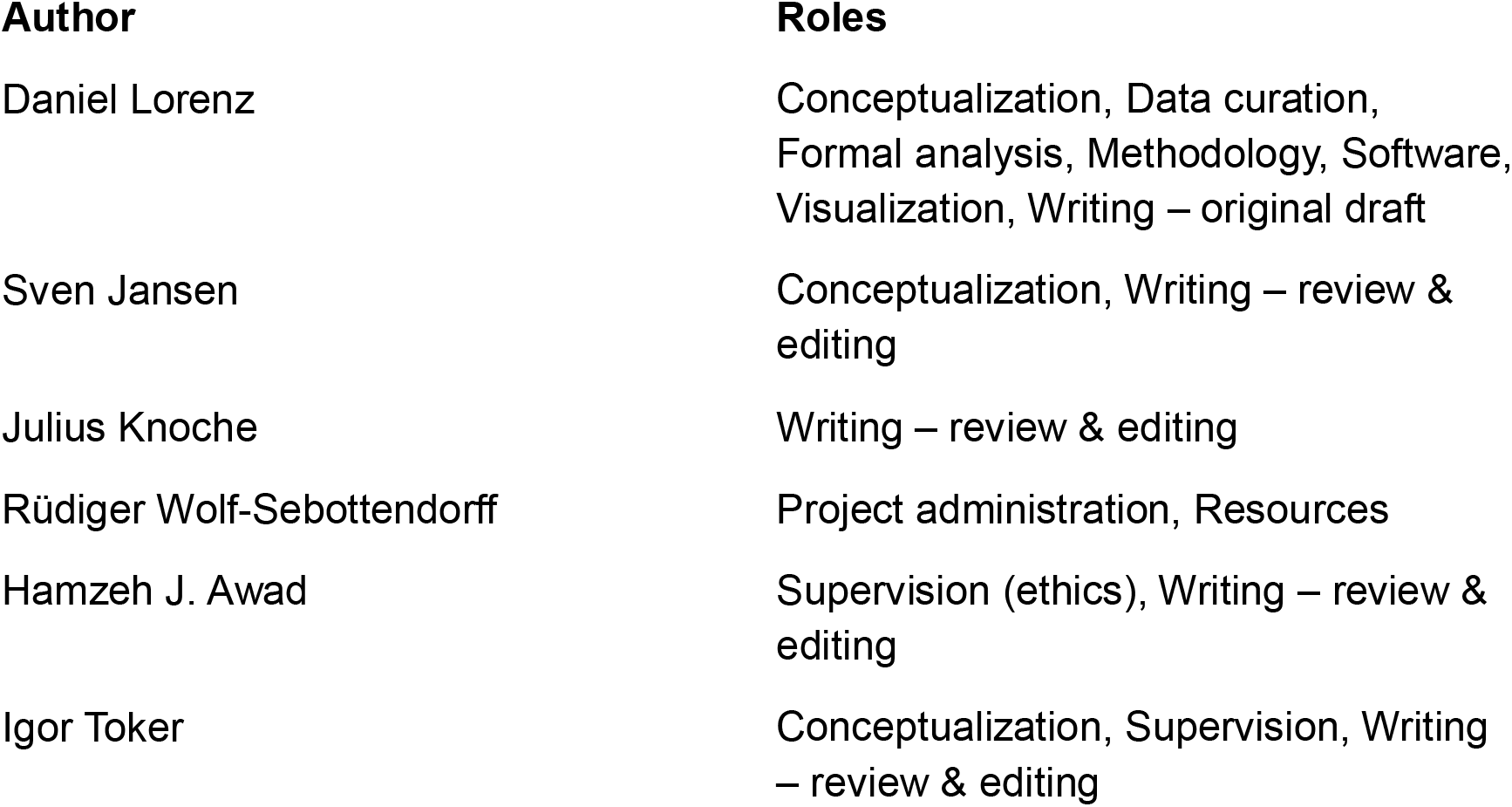

