## Supplemental Data 1 for "Adoption of Guided Structured Reporting in Routine Radiological Practice: A Six-Week Multi-Site Implementation Study in the UAE"

### Supplementary Table: Module-to-Modality Classification

System module names were mapped to imaging modality categories using keyword-based pattern matching. The classification rule was applied in the following order: (1) module names containing "CT" were classified as CT; (2) module names containing "MRT" or "MRI" were classified as MRI; (3) module names containing "MX" or "Mammo" (not already matched by rule 2) were classified as Mammography. Notably, "MammaMRT" (breast MRI) was correctly classified as MRI, not Mammography, because the "MRT" keyword match takes precedence.

| **Module Name** | **Modality** | **n (filtered dataset)** |
| --- | --- | --- |
| HeadCT | CT | 34 |
| ThoraxCT | CT | 16 |
| SpineCT | CT | 14 |
| AbdomenCT | CT | 10 |
| BoneCT | CT | 5 |
| AngiographyCT | CT | 3 |
| NeckCT | CT | 2 |
| **CT total** |  | **84** |
| HeadMRT | MRI | 7 |
| SpineMRT | MRI | 7 |
| PelvisMRT | MRI | 5 |
| AbdomenMRT | MRI | 4 |
| MammaMRT | MRI | 3 |
| KneeMRT | MRI | 1 |
| **MRI total** |  | **27** |
| MammaMX | Mammography | 15 |
| **Mammography total** |  | **15** |
| **Grand total** |  | **126** |
