## Supplemental Data 2 for "Adoption of Guided Structured Reporting in Routine Radiological Practice: A Six-Week Multi-Site Implementation Study in the UAE"

### Supplementary Material: Statistical Analyses

#### S1. Bootstrap Confidence Intervals for Weekly Medians

95% BCa bootstrap confidence intervals (10,000 resamples) for median active in-software reporting time (seconds) per modality and POC week.

### CT (n = 84)

| **Week** | **n** | **Median (s)** | **95% CI** |
| --- | --- | --- | --- |
| 1 | 12 | 130 | [83 – 204] |
| 2 | 15 | 166 | [92 – 271] |
| 3 | 21 | 111 | [52 – 170] |
| 5 | 25 | 49 | [30 – 93] |
| 6 | 11 | 56 | [28 – 124] |

##### MRI (n = 27)

| **Week** | **n** | **Median (s)** | **95% CI** |
| --- | --- | --- | --- |
| 1 | 8 | 383 | [256 – 521] |
| 2 | 4 | 368 | [238 – 515] |
| 3 | 6 | 101 | [79 – 138] |
| 5 | 7 | 122 | [64 – 168] |
| 6 | 2 | 60 | [48 – 71] |

##### Mammography (n = 15)

| **Week** | **n** | **Median (s)** | **95% CI** |
| --- | --- | --- | --- |
| 1 | 5 | 126 | [72 – 554] |
| 2 | 6 | 188 | [44 – 340] |
| 3 | 3 | 60 | [50 – 209] |
| 6 | 1 | 46 | n/a (single observation) |

Note: No reports were recorded in week 4 across any modality. Mammography had no reports in week 5. Wide confidence intervals reflect small subgroup sizes.

#### S2. Spearman Rank Correlation (Trend Test)

Spearman rank correlation between report sequence number (chronological order) and active in-software reporting time, per modality.

| **Modality** | **n** | **Spearman rho** | **p-value** | **Interpretation** |
| --- | --- | --- | --- | --- |
| CT | 84 | -0.403 | <0.001 | Significant decreasing trend |
| MRI | 27 | -0.780 | <0.001 | Strong significant decreasing trend |
| Mammography | 15 | -0.550 | 0.034 | Significant decreasing trend |

All three modalities show a statistically significant monotonic decrease in reporting time over the sequence of reports.

#### S3. Mixed-Effects Model

A linear mixed-effects model was fitted with log-transformed active reporting time as the dependent variable, POC week (continuous) and modality as fixed effects, and a random intercept for radiologist.

**Model specification:** log(Active reporting time) ~ POC Week + Modality + (1 | Radiologist)

| **Parameter** | **Estimate** | **SE** | **z** | **p-value** | **95% CI** |
| --- | --- | --- | --- | --- | --- |
| Intercept | 5.339 | 0.196 | 27.26 | <0.001 | [4.955 – 5.723] |
| POC Week | -0.264 | 0.045 | -5.93 | <0.001 | [-0.352 – -0.177] |
| Modality: MRI (vs. CT) | 0.589 | 0.169 | 3.48 | <0.001 | [0.257 – 0.920] |
| Modality: Mammography (vs. CT) | -0.025 | 0.275 | -0.09 | 0.927 | [-0.564 – 0.514] |
| Random intercept variance | 0.042 | 0.110 | — | — | — |

**Interpretation:** Each additional week was associated with a 23.2% decrease in active reporting time (exp(-0.264) = 0.768), after adjusting for modality and between-radiologist variability. The week effect was highly significant (p < 0.001). MRI reports took significantly longer than CT reports; mammography did not differ significantly from CT.

**Model fit:** n = 126 observations, 7 groups (radiologists), REML estimation, model converged.

#### S4. Sensitivity Analyses

Five sensitivity analyses were conducted to assess the robustness of the observed temporal trend. For each variant, the total sample size, median reporting time per modality, and Spearman trend test results are reported.

| **Variant** | **n** | **CT median (s)** | **MRI median (s)** | **Mammo median (s)** | **Spearman CT** | **Spearman MRI** | **Spearman Mammo** |
| --- | --- | --- | --- | --- | --- | --- | --- |
| Baseline (publication filter) | 126 | 92 | 168 | 126 | -0.40 | -0.78 | -0.55* |
| (a) No 20 s lower bound | 156 | 56 | 115 | 66 | +0.02 n.s. | -0.21 n.s. | -0.18 n.s. |
| (b) 2×IQR instead of 1.5×IQR | 127 | 92 | 168 | 149 | -0.40 | -0.78 | -0.61* |
| (c) Without Radiologist A | 55 | 104 | 160 | 126 | -0.38 n.s. | -0.85 | -0.55* |
| (d) Without UAE-Home data | 88 | 128 | 179 | 126 | -0.17 n.s. | -0.79 | -0.55* |
| (e) Without first 5 reports per user | 92 | 80 | 118 | 149 | -0.33 | -0.64 | -0.45 n.s. |

Significance: \*\*\ *p < 0.001, \*\ *p < 0.01, \* p < 0.05, n.s. = not significant.

##### Interpretation

- **(b) 2×IQR:** Results are virtually identical to the baseline, confirming robustness of the outlier threshold.
- **(e) Without first 5 reports:** The trend persists for CT and MRI, suggesting that the observed reduction is not solely attributable to initial training effects.
- **(c) Without Radiologist A:** MRI and mammography trends persist; CT trend direction is maintained (rho = -0.38) but loses significance due to reduced sample size (n = 55).
- **(d) Without UAE-Home data:** MRI and mammography trends persist; CT trend weakens, reflecting the substantial contribution of Radiologist A's remote reports to the CT dataset.
- **(a) No lower bound:** Inclusion of very short reports (<20 s) introduces noise that obscures the trend across all modalities. This supports the rationale for excluding technically incomplete sessions.

Overall, the temporal trend is robust across most sensitivity variants, with expected loss of statistical power in reduced subsamples.

#### S5. Case-Mix Distribution by Week

Number of reports per module and POC week.

| **Module** | **Week 1** | **Week 2** | **Week 3** | **Week 5** | **Week 6** | **Total** |
| --- | --- | --- | --- | --- | --- | --- |
| HeadCT | 4 | 10 | 7 | 8 | 5 | 34 |
| ThoraxCT | 3 | 1 | 5 | 5 | 2 | 16 |
| SpineCT | 0 | 2 | 2 | 7 | 3 | 14 |
| AbdomenCT | 3 | 1 | 3 | 3 | 0 | 10 |
| BoneCT | 1 | 1 | 2 | 0 | 1 | 5 |
| AngiographyCT | 0 | 0 | 2 | 1 | 0 | 3 |
| NeckCT | 1 | 0 | 0 | 1 | 0 | 2 |
| HeadMRT | 2 | 0 | 1 | 4 | 0 | 7 |
| SpineMRT | 2 | 1 | 3 | 1 | 0 | 7 |
| PelvisMRT | 2 | 2 | 1 | 0 | 0 | 5 |
| AbdomenMRT | 0 | 1 | 0 | 2 | 1 | 4 |
| MammaMRT | 1 | 0 | 1 | 0 | 1 | 3 |
| KneeMRT | 1 | 0 | 0 | 0 | 0 | 1 |
| MammaMX | 5 | 6 | 3 | 0 | 1 | 15 |
| **Total** | **25** | **25** | **30** | **32** | **14** | **126** |

Note: No reports were recorded during week 4. The distribution of examination types across weeks shows no systematic shift toward less complex examinations in later weeks.

#### S6. Time-of-Day Distribution

| **Time of day** | **n** | **Median (s)** | **Week 1** | **Week 2** | **Week 3** | **Week 5** | **Week 6** |
| --- | --- | --- | --- | --- | --- | --- | --- |
| Morning (6–12) | 61 | 144 | 60% | 68% | 37% | 31% | 57% |
| Afternoon (12–18) | 45 | 111 | 40% | 32% | 43% | 25% | 43% |
| Evening (18–24) | 15 | 36 | 0% | 0% | 7% | 41% | 0% |
| Night (0–6) | 5 | 45 | 0% | 0% | 13% | 3% | 0% |

Note: Week 5 had a substantial proportion of evening reports (41%), predominantly from a single radiologist using the system remotely from home. Evening and night reports were associated with shorter active reporting times, which may partly explain the lower medians observed in later weeks.

#### S7. Adoption Metrics

Three proxy indicators of workflow adoption were derived from system logs: (1) **Completion rate** — proportion of sessions where at least one structured report was generated (Amount of status 'Generate' ≥ 1); (2) **Export rate** — proportion of sessions where the report was exported or copied to clipboard for integration into the clinical system (Amount of status 'Export/CopyToClipboard' ≥ 1); (3) **Report finalization rate** — proportion of sessions closed on the final reporting page ('R'), indicating that the radiologist completed the full structured reporting workflow rather than abandoning mid-process.

##### Overall (n = 126)

| **Metric** | **n** | **%** |
| --- | --- | --- |
| Completion rate (Generate ≥ 1) | 126 | 100.0 |
| Export rate (Export/Copy ≥ 1) | 91 | 72.2 |
| Report finalization rate (Closed on page = R) | 113 | 89.7 |
| Early exit (Closed on page ≠ R) | 13 | 10.3 |

##### By POC Week

| **Week** | **n** | **Completion rate** | **Export rate** | **Finalization rate** |
| --- | --- | --- | --- | --- |
| 1 | 25 | 100% | 32% | 80% |
| 2 | 25 | 100% | 60% | 80% |
| 3 | 30 | 100% | 87% | 100% |
| 5 | 32 | 100% | 94% | 91% |
| 6 | 14 | 100% | 86% | 100% |

The export rate increased markedly from 32% in week 1 to 94% in week 5, suggesting growing integration of structured reports into the clinical workflow over the course of the POC.

##### By Radiologist

| **Radiologist** | **n** | **Completion rate** | **Export rate** | **Finalization rate** |
| --- | --- | --- | --- | --- |
| Radiologist A | 71 | 100% | 83% | 92% |
| Radiologist C | 14 | 100% | 57% | 79% |
| Radiologist B | 13 | 100% | 100% | 92% |
| Radiologist E | 9 | 100% | 33% | 100% |
| Radiologist F | 8 | 100% | 75% | 88% |
| Radiologist D | 7 | 100% | 14% | 86% |
| Radiologist G | 4 | 100% | 25% | 75% |

Note: Export rate variation between radiologists likely reflects differences in local workflow integration (e.g., availability of clipboard import in the PACS/RIS system) rather than differences in adoption intent. Radiologists D and G had low report volumes (n ≤ 7), limiting interpretability of their individual rates.
