## Supplemental Data 3 for "Adoption of Guided Structured Reporting in Routine Radiological Practice: A Six-Week Multi-Site Implementation Study in the UAE"

### STROBE Checklist for Observational Studies

This checklist follows the STROBE Statement (Strengthening the Reporting of Observational Studies in Epidemiology) for cohort, case-control, and cross-sectional studies. Reference: von Elm E, et al. The Strengthening the Reporting of Observational Studies in Epidemiology (STROBE) statement. Lancet 2007; 370: 1453–57.

| **#** | **Item** | **Recommendation** | **Reported** | **Location in manuscript** |
| --- | --- | --- | --- | --- |
| **Title and abstract** |  |  |  |  |
| 1a | Title | Indicate the study's design with a commonly used term | Yes | Title |
| 1b | Abstract | Provide an informative and balanced summary | Yes | Abstract (structured: Background/Objective/Methods/Results/Conclusions) |
| **Introduction** |  |  |  |  |
| 2 | Background/rationale | Explain the scientific background and rationale | Yes | Introduction, paragraphs 1–6 |
| 3 | Objectives | State specific objectives, including any pre-specified hypotheses | Yes | Introduction, final paragraph; Objective in abstract |
| **Methods** |  |  |  |  |
| 4 | Study design | Present key elements of study design early in the paper | Yes | Methods → Study Design ("retrospective observational case study") |
| 5 | Setting | Describe the setting, locations, and relevant dates | Yes | Methods → Setting and Participants (two hospitals, Pure Health, Abu Dhabi; Oct–Dec 2025) |
| 6a | Participants | Give eligibility criteria, sources, and methods of selection | Yes | Methods → Setting and Participants; Data Sources and Data Processing |
| 7 | Variables | Clearly define all outcomes, exposures, predictors, potential confounders | Partially | Outcome Definition (active in-software reporting time defined); confounders discussed in Limitations |
| 8 | Data sources/measurement | Describe sources of data and methods of assessment for each variable | Yes | Methods → Data Sources and Data Processing; Outcome Definition |
| 9 | Bias | Describe any efforts to address potential sources of bias | Yes | Methods → Data Cleaning; Discussion → Strengths and Limitations |
| 10 | Study size | Explain how the study size was arrived at | Yes | Methods → Data Cleaning (audit trail: 189 raw → 126 after filtering) |
| 11 | Quantitative variables | Explain how quantitative variables were handled | Yes | Methods → Data Cleaning (20 s threshold, IQR outlier removal); Statistical Analysis |
| 12a | Statistical methods | Describe all statistical methods | Yes | Methods → Statistical Analysis (descriptive, bootstrap CI, Spearman, mixed-effects model) |
| 12b | Subgroups and interactions | Describe any methods for examining subgroups and interactions | Yes | Analyses stratified by modality; sensitivity analyses in supplement |
| 12c | Missing data | Explain how missing data were addressed | Yes | Methods → Data Sources ("Entries with missing or non-valid timestamps ... were excluded") |
| 12d | Loss to follow-up | Address any loss to follow-up (cohort studies) | N/A | Retrospective analysis of system logs; no follow-up |
| 12e | Sensitivity analyses | Describe any sensitivity analyses | Yes | Methods → Statistical Analysis; Supplement S4 (5 variants) |
| **Results** |  |  |  |  |
| 13a | Participants: numbers at each stage | Report numbers at each stage of the study | Yes | Results → Dataset Overview (189 raw, 126 included); audit trail in Methods |
| 13b | Non-participation | Give reasons for non-participation at each stage | Yes | Methods → Data Cleaning (16 test users, 37 <20 s, 7 outliers, 3 outside POC window) |
| 14a | Descriptive data: characteristics | Give characteristics of study participants | Partially | 7 radiologists, pseudonymised; modality distribution reported. Hospital-level volume not available. |
| 14b | Descriptive data: missing | Indicate number of participants with missing data | Yes | Implicit: all 126 records have complete data after cleaning |
| 15 | Outcome data | Report numbers of outcome events or summary measures | Yes | Results → Temporal Development (medians with IQR per modality per week); adoption metrics |
| 16a | Main results | Give unadjusted and adjusted estimates with CI | Yes | Spearman (unadjusted), mixed-effects model (adjusted for modality and user); bootstrap CI |
| 16b | Category boundaries | Report category boundaries when continuous variables are categorised | Yes | POC weeks defined; modality classification in Supplement |
| 16c | Relative/absolute measures | Consider translating estimates into meaningful clinical measures | Yes | Percentage reduction week 1→6; absolute values in seconds |
| 17 | Other analyses | Report other analyses done — e.g., subgroup, interaction, sensitivity | Yes | Sensitivity analyses (S4); time-of-day analysis (S6); case-mix (S5) |
| **Discussion** |  |  |  |  |
| 18 | Key results | Summarise key results with reference to study objectives | Yes | Discussion → Key Findings |
| 19 | Limitations | Discuss limitations, including sources of potential bias | Yes | Discussion → Strengths and Limitations (training confounding, time-of-day, sample size, metric scope) |
| 20 | Interpretation | Give a cautious overall interpretation considering objectives, limitations, and other evidence | Yes | Discussion → Interpretation in Context; Conclusion |
| 21 | Generalisability | Discuss the generalisability of the study results | Partially | Discussed in Limitations (pilot setting, limited users); hospital-level detail partially provided |
| **Other information** |  |  |  |  |
| 22 | Funding | Give the source of funding and the role of the funders | Pending | COI and funding statement to be added |
